# End-user feedback of rapid diagnostics in rural Kenya

**DOI:** 10.1101/2024.02.23.24302446

**Authors:** Jeffrey W. Beard, Daphne N. Pariser, Stephanie Monticelli, Benjamin L. Miller

**Author notes:** Corresponding Author (JB). These authors contributed equally to this work (Jeffrey W. Beard, Daphne N. Pariser). First authors (Jeffrey W. Beard, Daphne N. Pariser).

## Abstract

Underserved communities in low-resource countries are disproportionately impacted by communicable diseases when compared to those in developed countries. These communities have limited access to life saving diagnostic laboratory tests making it difficult to treat communicable diseases like SARS-CoV-2 and Human Immunodeficiency Virus (HIV). Rapid diagnostic tests, like the COVID-19 antigen (Ag) test, play a crucial role in underserved communities by enabling fast and inexpensive diagnosis in low-resource settings. Unfortunately, these rapid test platforms often lack the accuracy and precision of their laboratory-based analogs, resulting in a need for improved rapid diagnostics. The World Health Organization’s (WHO) ASSURED (Affordable, Sensitive, Specific, User-friendly, Rapid and robust, Equipment-free, and Deliverable to end-users) criteria are often referenced in the development of diagnostic tests. In this work, we aim to provide guidance to the “user-friendly” component of ASSURED through end-user surveys taken in rural Kenya. In these surveys, we examine the user-friendliness of two of the most commonly used rapid diagnostic tests, the COVID-19 Ag test and pregnancy test, by assessing participants’ familiarity with the tests, their opinion of test appearance, and the perceived complexity of the operator’s workflow. We also examine community acceptance and desire for a self-test for the highly stigmatized HIV. We intend these results to help guide developers of future rapid diagnostic tests intended for low-resource communities.

## Introduction

Proper disease diagnosis informs medical decisions that affect both patient care and public safety, but the ability for a patient to be diagnosed is dependent on the local healthcare infrastructure. A global imbalance in healthcare resources like diagnostic testing heavily favors developed countries, where high-end testing facilities capable of sensitive molecular tests are readily accessible.^1^ In contrast, developing nations’ healthcare systems are limited to primarily low-resource clinics,^2^ constraining testing capabilities to rapid diagnostic tests that often use lateral flow assay (LFA) technology.^3,4^ LFAs are paper-based tests that use a colorimetric indicator to distinguish the presence of an analyte in a biological sample; a home pregnancy test is the most common example of an LFA.^5^ Their compact size, paper-based layers, and relatively inexpensive reagents allow LFAs to be manufactured at scale and transported with minimal financial requirement, making them well-suited for regions with limited medical capability.

The World Health Organization (WHO) developed ASSURED (Affordable, Sensitive, Specific, User-friendly, Rapid and robust, Equipment-free, and Deliverable to end-users) as design criteria for developing diagnostics for low-resource areas. Rapid diagnostics such as LFAs adhere to most of the WHO’s ASSURED criteria and are a cornerstone of healthcare because they can bridge the gap of healthcare disparities by bringing robust and user-friendly diagnosis to patients in remote and urban areas alike, facilitating proper treatment of communicable diseases and reducing transmission.^6,7^ However, for a new or existing diagnostic to be useful to a community, the test must first be adopted and accepted by the community.^8^ Studies have shown that communities in low-resource areas can remain reluctant to adopt a rapid diagnostic even when it has been proven to work effectively,^9,10^ mandating a deeper understanding of community needs and beliefs prior to initiating the development of a new diagnostic. Further studies are needed to understand how to design rapid diagnostic tests that will meet the societal needs and expectations of the low-resource communities for which they are designed.

To address that need, we conducted surveys in two low-resource areas of Kenya, Ngowswani, and Nkoilale, to determine the local population’s familiarity with and perception of self-performed rapid diagnostic tests. We chose to use two of the most used LFAs as examples of self-tests for our surveys: the COVID-19 Antigen (Ag) test and two forms of pregnancy tests. The COVID-19 Ag test and pregnancy test were not only chosen for their worldwide usage but also because they provided an opportunity to ask questions about different assay formats and workflows. The pregnancy test provides two versions of the same test, digital and analog. The COVID-19 Ag test used in our study provides a more complex self-testing workflow than the pregnancy test, highlighting the potential complexities and utilities of an at-home diagnostic tool. Unlike the one-step pregnancy test, the COVID-19 Ag test is multi-step with a more invasive sample collection procedure. We anticipated that the complexity and invasiveness could deter end users from using the test, rendering it useless.

We also explored the acceptance and theoretical useability of a hypothetical self-performed Human Immunodeficiency Virus (HIV) test. Of the 39 million people living with HIV worldwide, approximately two-thirds live in low-resource areas.^11^ As a result of the new world goal for 95% of people living with HIV to know the status of their infection, the WHO estimates a demand of 29 million HIV self-tests with an estimated total cost of $180 million USD per year by 2025.^12^ Currently, HIV testing in low-resource settings is done using HIV Antibody (Ab) LFAs because they are inexpensive, accessible, and user-friendly. Unfortunately, these tests are only reliable during the late stages of an HIV infection,^13^ resulting in a high probability of false negative results. There is strong evidence that patient outcomes improve significantly with earlier detection of acute HIV,^14^ and acute HIV is a leading cause of HIV transmission.^15^ In 2022, only 86% of people living with HIV worldwide knew their status. Therefore, there is a need for improved rapid HIV tests that detect HIV at earlier stages than current HIV LFA Ab tests to reach the world goal of 95% of people aware of their HIV status.

Given the estimated increase in demand and need for improved rapid HIV tests, it is reasonable to predict an increase in research and development of HIV diagnostics. In the WHO’s estimation, Kenya is one of the top 10 countries that account for 70% of the world’s projected self-HIV testing,^12^ making it one of the top locations worldwide to survey HIV self-testing. We performed these surveys to better understand community acceptance of self-tests and the stigma around HIV testing to help guide future designs of rapid diagnostics for low-resource communities.

## Materials and methods

### Ethics statement

Research was approved by the University of Rochester RSRB (Research Study Review Board; Study 00007637), which made the determination that the research met federal and university criteria for exemption (exempt category 2i: tests, surveys, interviews, or observation (non-identifiable)). Formal consent was verbally obtained from each participant (described further in the *Pre-Survey Procedures* subsection).

### Study sites

Surveys were conducted over three days at two primary locations in Kenya, Olbolet Siana Hills Academy in Nkoilale and Destiny Shaper Primary School in Ngoswani. The survey sessions were held outdoors, on the respective school grounds. The research was conducted at two distinct locations with the intention of including a broader array of participants, thereby enriching the diversity of the population statistics in our study. On average, participants in the study walked ∼2-5 kilometers to the survey locations. These locations had previously been used as study sites by Humans for Education. In the area WiFi and electricity connectivity can be poor but Humans for Education allowed us to choose suitable sites with stronger connectivity points and strong connections with the community, allowing for a smooth working environment.

### Geographical and infrastructural context

Both sites are situated in rural areas and have limited access to electricity and Internet. To access the sites the team needed to access via paved and unpaved roads and by local guides for directions as GPS was not available.

### Demographic and Bias Consideration

Prior to conducting the study at each site, the demographics of each school were assessed, and we determined that both schools largely had similar numbers of students and income. This allowed us to rule out significant bias in terms of income disparity.

### Participant eligibility and recruitment

All participants were eligible if they were over the age of 18 and were able to provide consent for participation. Participants were compensated with 500 Kenyan shillings. Compensation was disbursed immediately after the survey through M-PESA (mobile payment; Pesa is the Swahili word for payment), a common cash app to which 100% of our participants had access.

Recruitment was done via Humans for Education, an international organization with a local presence in our study site areas. Humans for Education worked with local schools, their own staff, and the local leaders and government to disseminate information about the surveys prior to the study commencement. Participation was on a voluntary basis, with individuals arriving either singly or in groups, on a first-come-first-serve basis.

Our study aimed to recruit 100 total participants across both sites starting October 3, 2022, and ending October 5, 2022. By the end of our survey collection, we successfully surveyed 125 participants, with 75 from Olbolet Siana Hills Academy and 50 from Destiny Shaper Primary School. Our work with Humans for Education enabled us to connect with the community quickly and boosted our recruitment efforts. We sought to have a balanced representation of gender and age. With regard to age, we fully achieved this goal: our youngest participant was 20, and the oldest was 106. The gender distribution was less balanced and consisted of 10.4% men and 89.6% women. We discuss potential reasons for that later in the manuscript.

### On-site survey team

Our survey team consisted of Jeffrey Beard, Daphne Pariser, and two contracted interpreters. The interpreters were contracted to interpret the local Maasai language and Swahili to and from English to allow for communication between the survey team and participants. Both interpreters were local residents of greater Narok County. The translators had previously worked with Humans for Education on numerous surveys and had training in data collection and sensitivity.

### Pre-survey procedures

Before initiating the survey, prospective participants were briefed in groups. Interpreters translated the survey information sheet, elucidating the survey’s scope, as per the Institutional Review Board (IRB) regulations. This study received exemption status from the University of Rochester Medical Center’s Research Subjects Review Board (RSRB) office. Informed consent was secured from every participant before the survey commencement. Informed consent was verbal after the explanation of all the information about the survey as required by the exemption status. A researcher in the survey itself documented the consent.

Physical examples of digital and analog Clear Blue pregnancy tests, an Abbott COVID-19 Ag test, and a lancet were used as visual aids prior to participants taking the survey. These tests were chosen as representative examples to contextualize the survey questions. The COVID-19 Ag test and pregnancy tests were chosen for this study because they are self-performed tests commonly used throughout the world, including in these areas, and thus provided reliable insight into how common self-testing is in remote regions. The lancet visual was added to gain a better understanding of whether participants would be willing to self-administer at home. The use of analog versus digital test readouts provides a comparison of the same test with two different functional forms to understand which form is more desirable. Step-by-step instructions for the proper use of each test were described using the example tests to ensure participants would be able to answer all the survey questions. Test examples were demonstrated to groups of 1-10 participants at a time. No biological testing was conducted, and when any participants were concerned if they would be performing the tests themselves, it was explained that only surveys were being conducted.

### Data collection

Surveys were recorded using Google Forms. No personal identifying information was collected. Anticipating connectivity issues in the remote survey areas, 200 print versions of the survey were prepared as a backup. During moments of no internet access, 16 surveys were completed on paper and subsequently transcribed into Google Forms upon restoration of WIFI connectivity. Resource limitations dictated that only two participants could be surveyed at a time. Surveys were recorded on Google Forms by either Jeffrey Beard with assistance from an interpreter or recorded directly by one of the interpreters.

### Survey design and questions

The survey was constructed as a mixed-format survey, encompassing multiple-choice, image-based, and open-ended questions. Initial questions aimed to collect demographic data, including age, gender, and the number of children each respondent had. Respondents were asked about their prior experience with home diagnostic tests to better understand their experiences, specifically with pregnancy and COVID-19 tests. Those with prior experience were further asked to specify which tests they were familiar with. Using visual aids, participants were shown two types of pregnancy tests. They were asked about their preferences between the two and the reasons for their preferences. They were also asked about which result display method they preferred (analog or digital). Questions related to the perceived complexity of using a COVID-19 home test were also asked.

Additionally, respondents were asked about their disposal practices after test usage and their comfort level in using such a test on their own, rather than at a doctor’s office.

The survey explored participants’ willingness to pay for an HIV home test and their likelihood of using one if provided for free. Questions also delved into the comfort level of self-administering the test, specifically the act of pricking one’s finger, and the perceived utility of such a test in the community. The survey also probed for potential concerns or fears related to home-based HIV testing. Lastly, participants were given an opportunity to express their perceived need for similar tests for other diseases and to provide general feedback on how to improve the tests presented.

### Data analysis

All collected data were imported to statistical software (JMP Pro 17) for analysis. Any unanswered questions were omitted from the analysis. Non-responses by participants led to missing survey data for specific questions when the participant elected not to answer the survey question; however, no survey question had more than three missing data points. One of our survey questions, regarding payment for HIV tests, was only posed to the last 50 participants due to interpretation concerns, and thus the initial 75 participants were excluded from this analysis. Statistical significance was calculated using the one-sided Fisher’s Exact Test, where a = 0.05, p < 0.05 for *, p < 0.01 for **, and p < 0.001 for ***.

Qualitative data from the mixed-format surveys was coded based on response content and then organized by thematic analysis. For example, the question referring to the digital versus analog pregnancy test design, “Why do you prefer that test over the other?” was only broken into three categories: “Easy to Interpret,” “Cannot Read English,” and “Superior Technology,” because answers consistently fit these themes. The question, “Is there anything that scares you about taking an HIV test at home?” was broken into 18 categories to encompass the wide variety of responses.

### Limitations and barriers

First, the survey was conducted in only two primary locations within Kenya, which might not represent the broader rural or urban populations of the country. Therefore, we aim to expand our scope in future work by increasing the sample size and incorporating multiple locations. Printed surveys were used during Wi-Fi outages and therefore the transcription of these surveys into digital format may have introduced minor errors or discrepancies. In the future, we will implement a double-entry verification system, where two independent researchers input the data to ensure accuracy and consistency in the digital records. The recordings of the open-ended questions were translated by interpreters and thus could be subject to bias through interpretation. Due to the limited availability of local language interpreters within the region, an overlap between interpreters and survey participants who may have known each other was an unavoidable circumstance.

## Results

### Participant demographics

A total of 126 participants were surveyed in Kenya about home diagnostic testing (Fig 1a), with one participant removed from the analysis due to age requirement restrictions. Geographically there were 75 participants in Nkoilale and 50 participants in Ngowswani that participated in this study (Fig 1b). Participants were not asked about their residency, so a participant’s location is based solely on where they took the survey. Our participants ranged from 20 to greater than 70 years of age, with a mean of 33.6 and a median of 30 years (Fig 1c) and included a total of 112 women and 13 men (Fig 1d). It is unclear if the skew towards female participants was due to cultural reasons (e.g., women bringing their children to school), or if it had to do with the dissemination of information during recruitment. Participants had an average of 4 children, spanning from 0 to 10 children per participant (Fig 1e).

**Fig 1.**
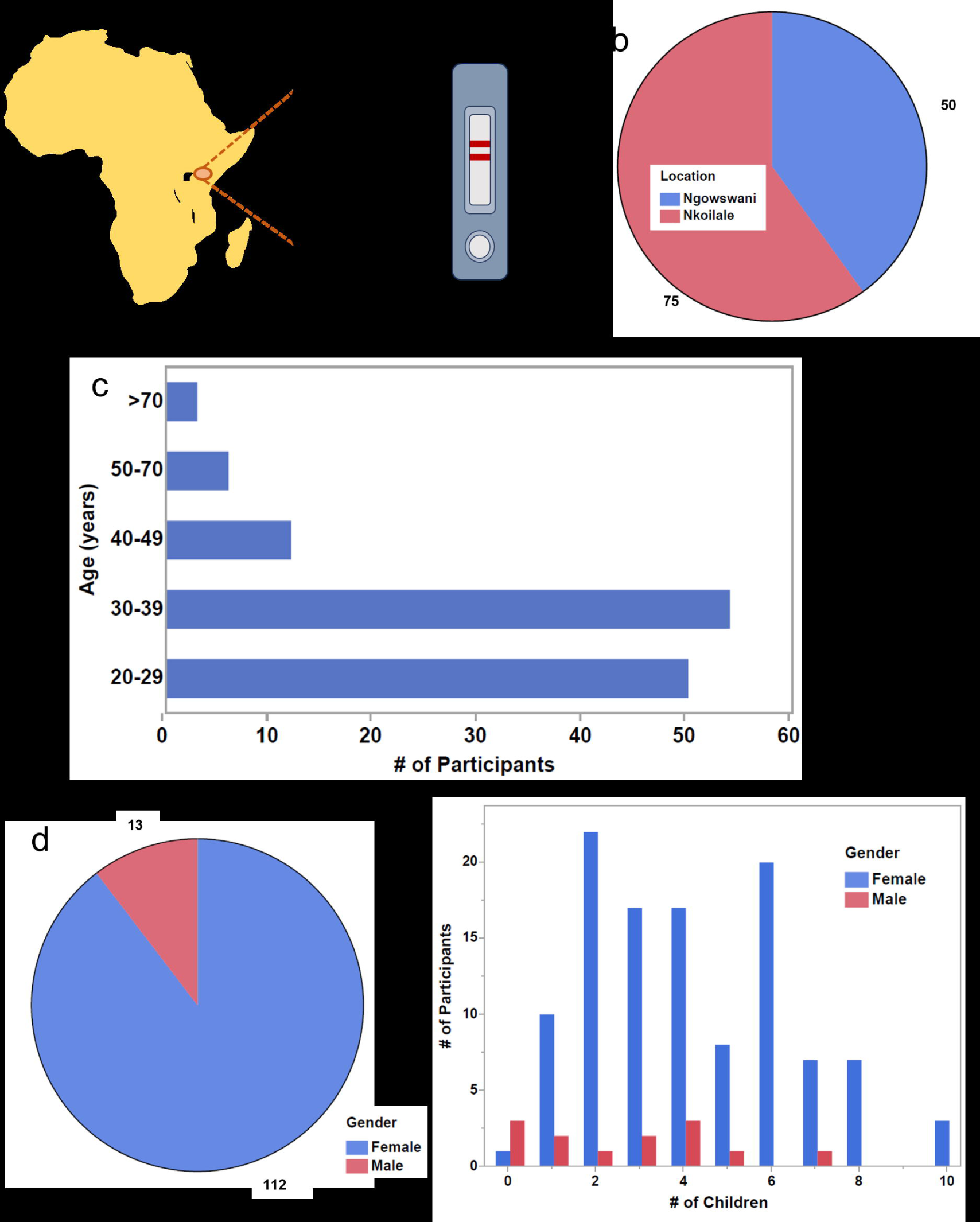
Survey demographics indicate that participants are primarily women from Nkoilale. a) Graphical abstract of rapid diagnostic surveys in Kenya. b) This pie chart indicates that the majority of participants are from Nkoilale, Kenya. c) The majority of participants are ages 20 - 40 years old. d) Our participants are broken down by gender in this pie chart revealing that they are primarily female. e) Participants categorized by gender and the number of children they reported having indicates participants have on average 2 - 6 children.

### Rapid diagnostic test familiarity

To assess participants’ familiarity with home diagnostic tests, they were asked to disclose if they had ever used a pregnancy test or COVID-19 Ag test. Both the pregnancy and COVID-19 Ag tests in question use the same lateral flow, paper-based technology that enables cost-effective diagnosis of a specific disease or medical condition at the point of care. Their paper-based platform is shared by other important, but less commonly used, rapid diagnostics such as HIV antibody tests and Malaria Ag tests. Over 85% of participants reported being familiar with at least one of the home tests demonstrated, with no significant difference between the male and female participants (Figs 2a and 2b). A large majority of participants (79.2%) were familiar with the pregnancy test while a minority (30.4%) were familiar with the COVID-19 test. Less than a quarter of participants were familiar with both (Fig 2c). Participants from the Nkoilale survey location were more likely to be familiar with the COVID-19 test than participants in the Ngowswani location (Fig 2d), while there was no significance between the two locations regarding familiarity with the pregnancy test (Fig 2e).

**Fig 2.**
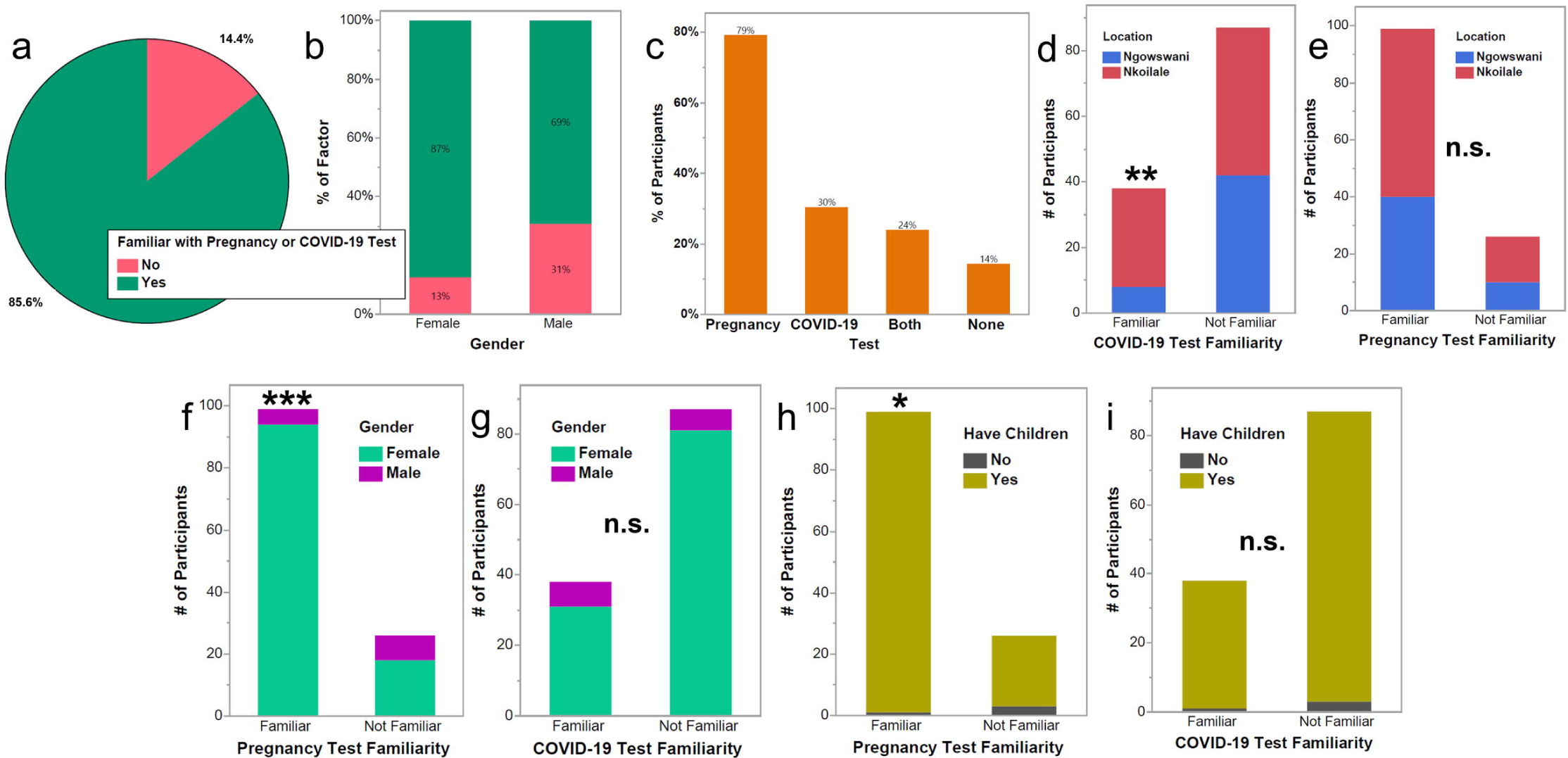
Participants are more familiar with pregnancy home diagnostic tests, especially when they have children. a) Pie chart showing 14.4% of participants were not familiar with either the pregnancy test or COVID-19 test, and b) percentage of participants by gender familiar with at least one of the tests. There is no statistical significance between gender and familiarity with at least one test. c) Percent of participants familiar with the pregnancy test (79%), COVID-19 test (30%), both tests (24%), or none of the tests (14%). d) and e) The number of participants familiar with the COVID-19 test (d) or the pregnancy test (e) by location.

Participants in the Nkoilale location were significantly more likely to be familiar with the COVID-19 test than those from the Ngowswani location. f) and g) The number of participants familiar with the pregnancy test (f) or COVID-19 test (g) by gender. Surveyed women were significantly more likely to be familiar with the pregnancy test than men. h) and i) The number of participants familiar with the pregnancy test (h) or COVID-19 test (i) by whether the participant has children. Participants with children were significantly more likely to be familiar with the pregnancy test than those without children.

We found that women were significantly more likely to be familiar with the pregnancy test than men (Fig 2f). Men were more familiar with the COVID-19 test (Fig 2g), trending towards statistical relevance (p-value= 0.056) in the future we look to increase the number of male participants. The skew towards pregnancy test familiarity in Fig 2c may be due, at least in part, to the bias of female participant turnout over male. We also found that participants with children were more likely to be familiar with the pregnancy test than those without (Fig 2h), and the factor of children had no relevance towards familiarity to the COVID-19 test (Fig 2i). Age did not have statistical relevance towards test familiarity when participants were categorized into greater than or less than 30 years of age.

### Test design preferences

When asked to choose between the digital and analog pregnancy tests, nearly 80% of all participants chose the digital test (Fig 3a). When asked why participants chose the digital over the analog tests, the responses consistently referred to either the simple interpretation of the digital display or the idea that the digital test contained superior technology. Only a small fraction of the participants thought the pregnancy tests could be improved, with the top response being the inclusion of an additional test to come with the first to confirm the diagnostic results (Fig 3b).

**Fig 3.**
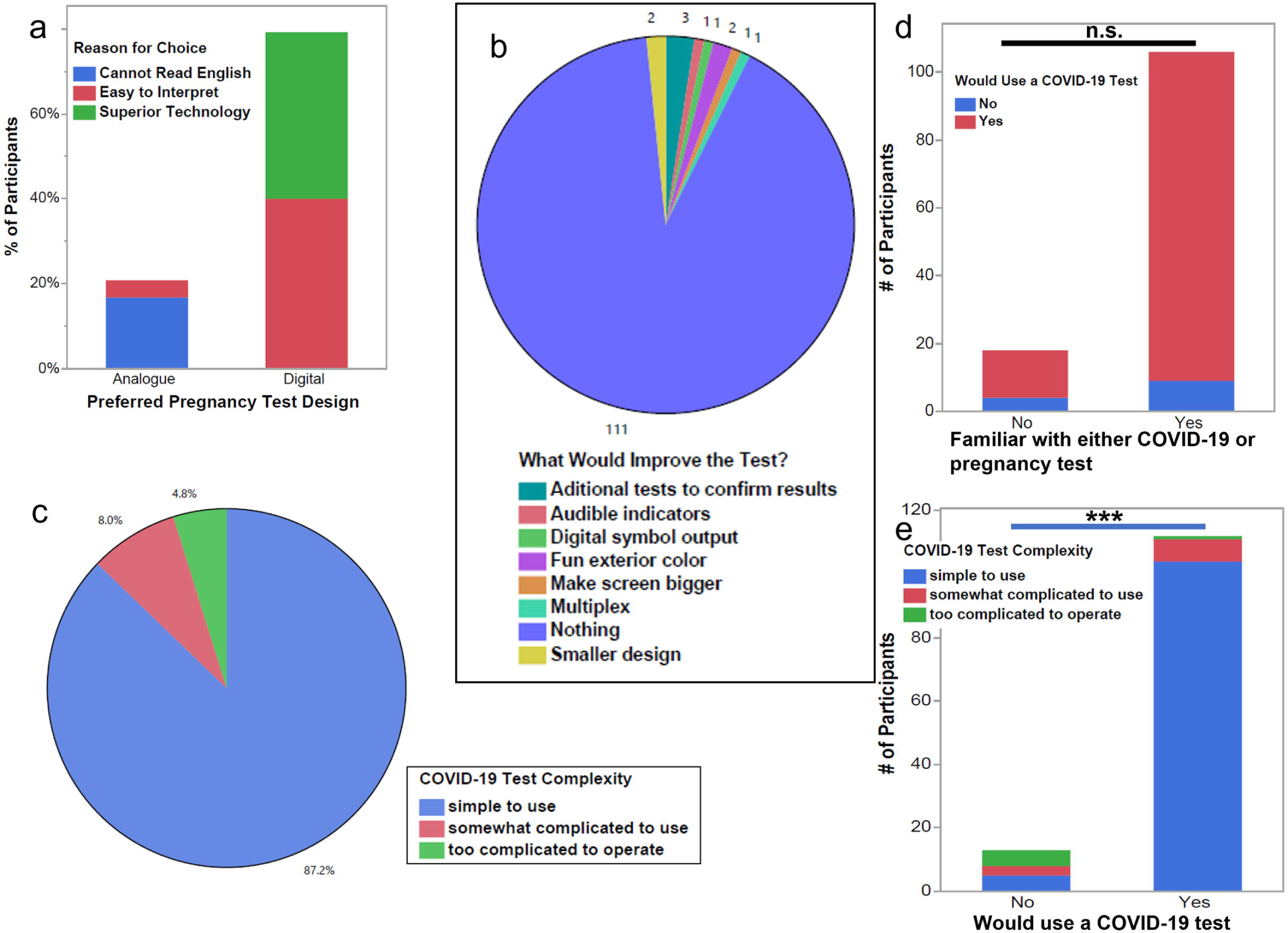
User opinions of pregnancy and COVID-19 tests. a) Percent of participants who preferred the analog or digital pregnancy test and the categorized reasons for choosing that test. The majority of participants who chose the analog test reported not being able to read English. b) Pie chart showing participant recommendations to improve the pregnancy home test. Most participants provided no recommendations. c) Pie chart showing percentages of total responses rating the complexity of the COVID-19 test workflow from simple (87.2%), somewhat complicated (8%), and too complicated to operate (4.8%). d) A bar graph showing that familiarity with the COVID-19 or pregnancy test does not correlate with whether a participant would use a COVID-19 test. e) A bar graph showing that participants who viewed the COVID-19 test as “simple to use” were statistically more likely to use a COVID-19 test.

After a demonstration of the Abbott COVID-19 Ag test, the participants were asked to rate the user-friendliness of the test in terms of workflow complexity. Over 87% of the participants reported the COVID-19 test as being simple to use, and nearly 13% reported the test as somewhat to extremely complicated to use (Fig 3c). Prior exposure to COVID-19 or pregnancy tests did not have an impact on whether a participant would use a free version of the COVID-19 test (Fig 3d). However, participants who rated the complexity of the COVID-19 test as simple to use were significantly more likely to be willing to use such a test (Fig 3e).

### Survey questions about HIV self-testing

Although 100% of survey participants believed an HIV test would benefit their community (Fig 4a), 4% of participants reported that they would not use a free HIV test (Fig 4b). The reasons given for why the 4% of participants would not use the HIV test included: only doctors can administer tests, being afraid of the results when alone, and being anxious about not using the test correctly. The 13.9% of participants who reported not being willing to use a lancet to draw their own blood (Fig 4c) also compose over 10% of the participants who said they would use a home HIV test (Fig 4d).

**Fig 4.**
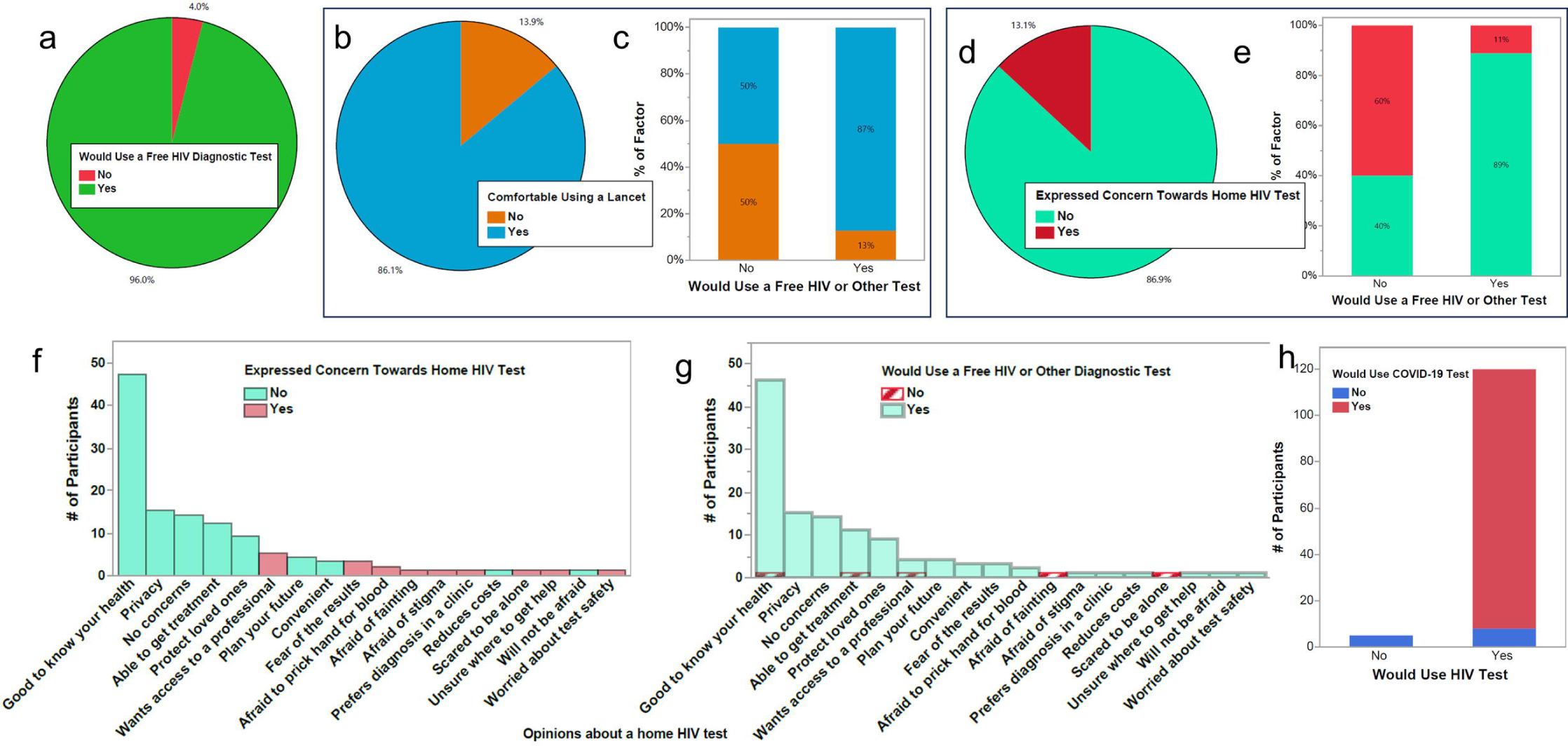
Participant reactions towards a home HIV test. a) Pie chart showing 96% of participants would use a free HIV test. b) Pie chart showing 86.1% of participants would be comfortable using a lancet. c) Bar graph showing 13% and 50% of participants who said they would or would not use a free HIV test, respectively, also said they would not be comfortable using a lancet. d) Pie chart showing 86.9% of participants had no concerns about using a home HIV test. c) Bar graph showing that 11% and 60% of participants who said they would or would not use a free HIV test, respectively, also expressed concerns about a home HIV test. f) Participant opinions about a home HIV test and whether the participants expressed concerns about a home HIV test. g) Participant opinions about a home HIV test and whether the participants would use a free HIV test. h) Number of participants who would or would not use an HIV test compared with if they would or would not use a COVID-19 test.

Only 13% of participants expressed concerns about taking a home HIV test (Fig 4e). As shown in Fig 4f, these individuals expressing concerns also make up 11% of the participants who were willing to use a home HIV test. Fig 4g shows the wide range of feedback participants gave as to why or why not they felt a home HIV test would be good for their communities. Participants’ top concerns (shown as red bars) were not having immediate access to a professional for counseling or healthcare, facing the results of the test, and pricking their own finger for the necessary blood sample. The majority of participants who expressed these top three concerns were also participants who said they would use a home HIV test if provided for free (Fig 4h), and more participants were willing to use the home HIV test over the home COVID-19 test (Fig 4i).

### Economic and environmental considerations

Fig 5a shows the breakdown of 50 participants who were asked how much they would pay for a rapid HIV test, where 86% said they would not pay more than 100 KSH (about $0.67 USD as of October 2022) for an HIV test. From those 50 respondents, 75% of the people who said they would not use a rapid HIV test also said they would pay money for an HIV test (Fig 5b).

**Fig 5.**
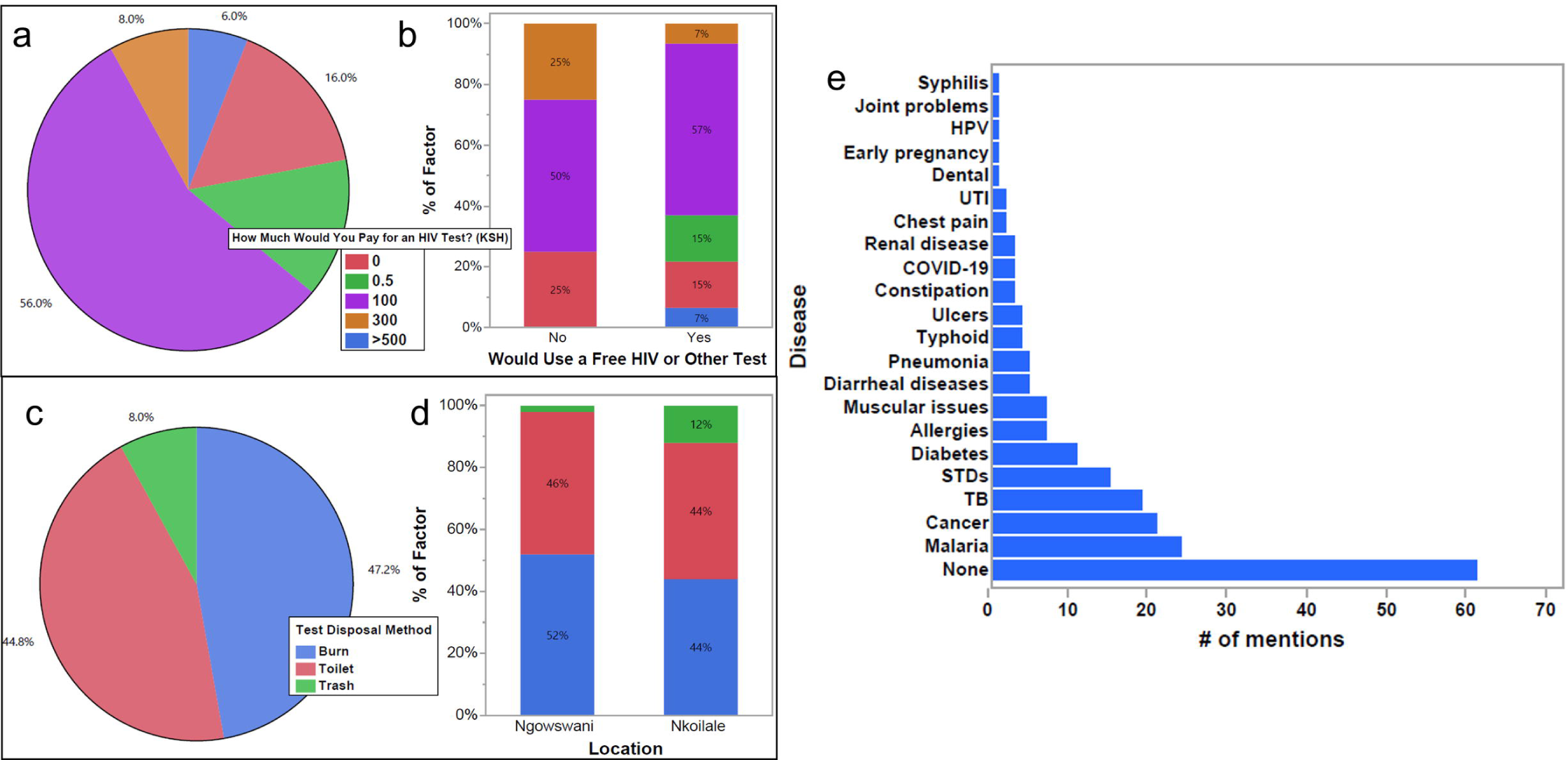
Economic and environmental considerations. a) Pie chart showing the percentage of respondents who reported how much they would pay for an HIV test in Kenyan shillings (KSH). 16% said 0 KSH, 56% said 100 KSH, 8% said 300 KSH, and 6% said >500 KSH. b) A bar graph showing participants grouped by who would or would not use an HIV test and broken down by how much they would pay for an HIV test by percentage of the group. c) Pie chart showing the percentage of respondents categorized by how they would dispose of a home diagnostic test. Three responses were given: burn (47.2%), throw in the toilet (44.8%), or throw in the trash (8.0%) d) A bar graph showing participants grouped by location and broken down into how they would dispose of a test by the percentage of the group. e) At the end of each survey, participants were asked to provide examples of any other diagnostic tests they felt would benefit their communities. Their responses were grouped and plotted as a bar graph by the number of times each disease was mentioned.

When asked how participants would dispose of a used COVID-19 test, one of three answers was provided: burn, throw away in the trash bin, or throw away in the toilet, making up 47.2%, 8%, and 44.8% of participant responses, respectively (Fig 5c). These responses trended the same across both survey locations (Fig 5d), highlighting that environmental and biological safety should be considered when designing future diagnostics for low-resource settings.

At the end of the survey, we asked participants to list other diseases for which a self-test would be useful. The purpose of this open-ended question was to understand what ailments were affecting locals. Fig 5a shows that approximately 50% of participants listed diseases they thought would be useful for self-testing, which included malaria, cancers, tuberculosis, sexually transmitted infections, and diabetes.

## Discussion

To better understand the level of exposure to and use of LFAs among residents in low-resource communities, as well as to evaluate the potential acceptance of a rapid HIV self-test in these communities, we conducted surveys in Narok County, Kenya. This study is crucial to the development of effective rapid HIV diagnostics because it encourages input from the end user. Since the pregnancy test is the most widely used home test in the world, it is not surprising that most participants, male and female alike, were familiar with how it worked. However, given the scale and high global impact of the COVID-19 pandemic, it is surprising that only 30% of participants were familiar with the COVID-19 Ag test.

The survey site at Nkoilale was situated nearer to the primary government COVID-19 testing facility compared to the Ngoswani site. This proximity may account for the higher awareness of COVID-19 testing among Nkoilale participants versus those from Ngoswani. However, such a correlation between location and awareness did not apply to pregnancy testing. It’s worth noting that a temporary COVID-19 testing outpost, positioned roughly a ten-minute drive from Nkoilale and thirty minutes from Ngoswani, was operational for a period. Despite this, the majority of survey participants, due to the cost barrier, did not utilize this service, and consequently, attendance was low. This temporary site has since been discontinued, and currently, the closest testing center is an hour’s drive away for both locations. The high global demand and low supply during the 2020 pandemic is a possible explanation for the low percentage of familiarity with the COVID-19 test. During the heart of the pandemic, rapid COVID-19 diagnostics were primarily funneled to wealthier countries, leaving low-resource communities deficient in supplies.^16^ Additionally, for the majority of participants there were no vaccination sites within walking distance and even though the vaccines were delivered for free transportation to a nearby clinic was too expensive.

Even though both pregnancy tests use the same LFA technology under their respective casing, there was an apparent increase in confidence in the digital pregnancy test over the analog. The digital pregnancy test contains electronics to analyze the test and provide a digital readout of the test results. In contrast, the analog test contains no electronics, and the results must be interpreted by the user. The main advantage of the digital pregnancy test was that it took away the ambiguity of the analog test, making it easier to interpret, which is crucial in communities with low literacy rates or where multiple languages and local dialects are spoken. According to the World Bank, the literacy rate in Kenya in 2021 was 83%.^17^ This is consistent with the percentage of participants who chose the analog test over the digital one for literacy reasons, which was also verified in our study. Of the 20% of participants who chose the analog test over the digital, all expressed concerns about being able to interpret the digital test display, with many explicitly saying they could not read the English words displayed on the digital screen. Although this survey does not definitively determine what test output would be universally favored, it does demonstrate the importance of being able to interpret the test results. An ideal diagnostic test will be easily interpreted by the end user, even if they cannot read English.

Even though the digital test display was favored over the analog, it is clear environmental impacts should be considered when designing medical equipment for low-resource communities. Improper burning of the test consumables (i.e. the rapid diagnostic tests themselves) results in the release of carcinogens,^18^ and even throwing the tests into the trash in low-resource settings often ends with improper disposal or burning of the trash.^19^ The addition of unnecessary electronics should be avoided at all costs to reduce the impact of improper waste disposal. Industry standards like nitrocellulose can be harmful to the environment, so biodegradable components such as compostable cardboard packaging^20^ and cellulose flow strips^21,22^ should be considered when choosing materials for new rapid diagnostic tests.

Aside from protecting the environment, unnecessary electronics should be excluded from development for financial reasons. Most diagnostics for low-income countries are supplemented by non-government organizations. However, the current average price for a rapid HIV test of $5.46 USD^11^ should be maintained or lowered even as the test technology is improved. This is of particular importance because 86% of participants said they would not pay more than 100 KSH (approximately $0.66 USD) for an HIV test.

Although most participants said they would use a COVID-19 or HIV test if provided to them for free, it is interesting that 8 participants were willing to use the hypothetical home HIV test over the home COVID-19 test. It is likely the difference in test useability lies in the biological sample. The collection of biological samples for the COVID-19 test and HIV test were the primary difference between the tests, where the COVID-19 test required the addition of a nasal swab sample and the hypothetical HIV test required the addition of a blood sample retrieved by lancing a finger. The 6.4% of participants who said they would use an HIV test but not a COVID-19 test, also said that they would be willing to draw their own blood using a lancet for the HIV test, indicating that using a nasal swab is less appealing than using a lancet.

Aside from the method of sample acquisition, many participants were concerned about not having access to a counselor or medical professional when using a home HIV test; even participants who said they would use the home HIV test had these concerns. Although this concern is not directly joined to the acceptability of a rapid diagnostic test, it should be considered as an associated problem. If an individual is going to use a diagnostic test for a life-threatening disease, there is a moral obligation for the developers of the diagnostic test to provide some linking service to healthcare professionals for follow-up support.

There is a surplus of research to develop novel diagnostics that can operate in low-resource conditions. This work strives to provide an understanding of how current diagnostics are viewed in low-resource regions to guide the development of these diagnostics. It is crucial for developers to design future diagnostic tests in a manner that the end users will accept, and this survey contributes towards that end. Our future work will include investigating the acquisition and distribution of rapid diagnostics in low-resource regions to understand why there is variability in familiarity with common diagnostics such as the COVID-19 Ag test. We also aim to produce an HIV self-test that addresses the concerns described earlier. The absence of critical medical supplies, such as diagnostics for pervasive diseases like COVID-19 and HIV, is common in low-resource facilities,^23^ and a deeper understanding of distribution methods is necessary to increase the availability of healthcare in underserved regions.

## Data Availability

The data that support the findings of this study are available in the supplementary material of this article.

## Acknowledgments

We graciously thank our hosts in the Maasai Mara region of Kenya and everyone who participated in our survey. We would also like to thank our interpreters for their hard work and the staff of Humans for Education.

